# SARS-CoV-2 Seroprevalence Among Parturient Women

**DOI:** 10.1101/2020.07.08.20149179

**Authors:** Dustin D. Flannery, Sigrid Gouma, Miren B. Dhudasia, Sagori Mukhopadhyay, Madeline R. Pfeifer, Emily C. Woodford, Jeffrey S. Gerber, Claudia P. Arevalo, Marcus J. Bolton, Madison E. Weirick, Eileen C. Goodwin, Elizabeth M. Anderson, Allison R. Greenplate, Justin Kim, Nicholas Han, Ajinkya Pattekar, Jeanette Dougherty, Oliva Kuthuru, Divij Mathew, Amy E. Baxter, Laura A. Vella, JoEllen Weaver, Anurag Verma, Rita Leite, Jeffrey S. Morris, Daniel J. Rader, Michal A. Elovitz, E. John Wherry, Karen M. Puopolo, Scott E. Hensley

**Affiliations:** Division of Neonatology, Children’s Hospital of Philadelphia, Philadelphia, PA; Department of Pediatrics, University of Pennsylvania Perelman School of Medicine, Philadelphia, PA; Center for Pediatric Clinical Effectiveness, Children’s Hospital of Philadelphia, Philadelphia, PA; Department of Microbiology, University of Pennsylvania Perelman School of Medicine, Philadelphia, PA; Division of Infectious Diseases, Children’s Hospital of Philadelphia, Philadelphia, PA; Institute for Immunology, University of Pennsylvania Perelman School of Medicine, Philadelphia, PA; Department of Systems Pharmacology and Translational Therapeutics, University of Pennsylvania, Philadelphia, PA; Division of Gastroenterology, Department of Medicine, University of Pennsylvania Perelman School of Medicine, Philadelphia, PA; Institute for Translational Medicine and Therapeutics, University of Pennsylvania Perelman School of Medicine, Philadelphia, PA; Departments of Genetics and Medicine, Perelman School of Medicine, University of Pennsylvania, Philadelphia, PA; Maternal and Child Health Research Center, Department of Obstetrics and Gynecology, University of Pennsylvania Perelman School of Medicine, Philadelphia, PA; Department of Biostatistics Epidemiology and Informatics, University of Pennsylvania, Philadelphia, PA

## Abstract

Limited data are available for pregnant women affected by SARS-CoV-2. Serological tests are critically important to determine exposure and immunity to SARS-CoV-2 within both individuals and populations. We completed SARS-CoV-2 serological testing of 1,293 parturient women at two centers in Philadelphia from April 4 to June 3, 2020. We tested 834 pre-pandemic samples collected in 2019 and 15 samples from COVID-19 recovered donors to validate our assay, which has a ∼1% false positive rate. We found 80/1,293 (6.2%) of parturient women possessed IgG and/or IgM SARS-CoV-2-specific antibodies. We found race/ethnicity differences in seroprevalence rates, with higher rates in Black/non-Hispanic and Hispanic/Latino women. Of the 72 seropositive women who also received nasopharyngeal polymerase chain reaction testing during pregnancy, 46 (64%) were positive. Continued serologic surveillance among pregnant women may inform perinatal clinical practices and can potentially be used to estimate seroprevalence within the community.

**One Sentence Summary:** Six percent of pregnant women delivering from April 4 to June 3, 2020 had serological evidence of exposure to SARS-CoV-2 with notable race/ethnicity differences in seroprevalence rates.

---

Severe acute respiratory syndrome coronavirus 2 (SARS-CoV-2) can cause serious disease in adult populations, particularly in those with underlying health conditions (1). SARS-CoV-2 serological tests are important for determining immunity within individuals and populations (2). However, many commercial tests have high false positive rates and therefore cannot be used to accurately estimate seroprevalence in populations with relatively low levels of exposures (3,4). Serological tests are especially important for vulnerable populations such as pregnant women, because immune status has implications for management of both the pregnant woman and the newborn. Admission to the hospital for delivery is one of the few instances in which otherwise healthy individuals are consistently interacting with the medical system, and therefore provides an opportunity for population surveillance of SARS-CoV-2 serology.

We performed a prospective cohort study of pregnant women presenting for delivery from April 4 to June 3, 2020 at two academic birth hospitals in Philadelphia, Pennsylvania. Both hospitals are active clinical and research centers affiliated with the University of Pennsylvania, and combined represent 50% of live births in Philadelphia (5). Discarded maternal sera from delivery admission were collected, deidentified, and tested by enzyme-linked immunosorbent assay (ELISA) for SARS-CoV-2 immunoglobulin G (IgG) and immunoglobulin M (IgM) antibodies to the spike receptor binding domain (RBD) antigen.

Demographics and clinical characteristics of the women are shown in **Table 1**. Most serum specimens were derived from women living in areas within or immediately bordering the city of Philadelphia (**Figure 1**). Symptomatic pregnant women and those with known risk factors underwent SARS-CoV-2 nasopharyngeal (NP) nucleic acid polymerase chain reaction (PCR) testing from April 4-12, 2020; universal PCR testing was recommended for all pregnant women presenting for delivery starting April 13, 2020. Of 1,620 women who delivered during the study period, 1,293 (80%) had available discarded serum specimens and were included in the analysis.

**Table 1.**
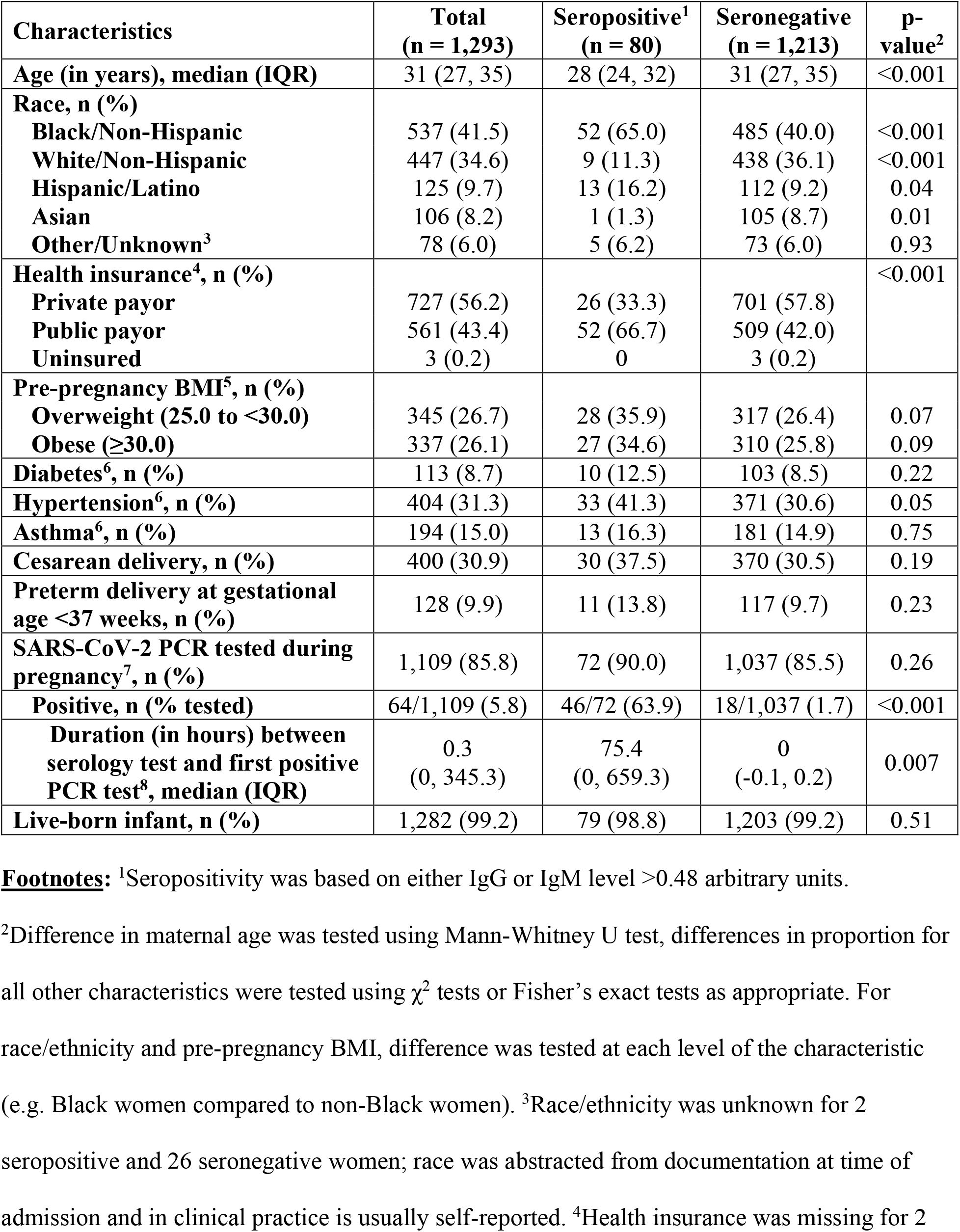
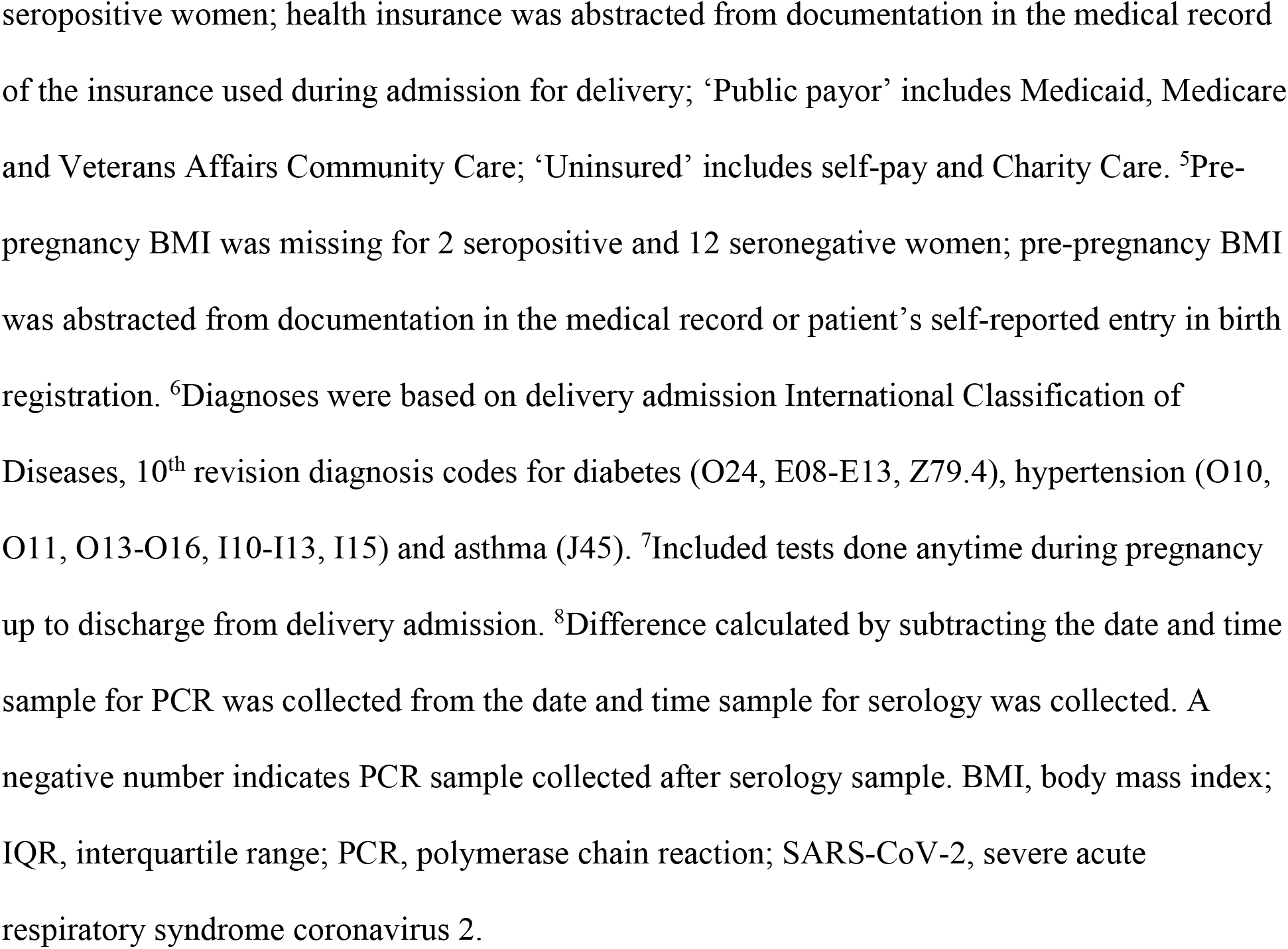
Demographics and Clinical Characteristics of the Study Cohort.

| Characteristics | Total<br>(n = 1,293) | Seropositive<sup>1</sup><br>(n = 80) | Seronegative<br>(n = 1,213) | p-value<sup>2</sup> |
| --- | --- | --- | --- | --- |
| Age (in years), median (IQR) | 31 (27, 35) | 28 (24, 32) | 31 (27, 35) | <0.001 |
| Race, n (%) |  |  |  |  |
| Black/Non-Hispanic | 537 (41.5) | 52 (65.0) | 485 (40.0) | <0.001 |
| White/Non-Hispanic | 447 (34.6) | 9 (11.3) | 438 (36.1) | <0.001 |
| Hispanic/Latino | 125 (9.7) | 13 (16.2) | 112 (9.2) | 0.04 |
| Asian | 106 (8.2) | 1 (1.3) | 105 (8.7) | 0.01 |
| Other/Unknown<sup>3</sup> | 78 (6.0) | 5 (6.2) | 73 (6.0) | 0.93 |
| Health insurance<sup>4</sup>, n (%) |  |  |  | <0.001 |
| Private payor | 727 (56.2) | 26 (33.3) | 701 (57.8) |  |
| Public payor | 561 (43.4) | 52 (66.7) | 509 (42.0) |  |
| Uninsured | 3 (0.2) | 0 | 3 (0.2) |  |
| Pre-pregnancy BMI<sup>5</sup>, n (%) |  |  |  |  |
| Overweight (25.0 to <30.0) | 345 (26.7) | 28 (35.9) | 317 (26.4) | 0.07 |
| Obese (≥30.0) | 337 (26.1) | 27 (34.6) | 310 (25.8) | 0.09 |
| Diabetes<sup>6</sup>, n (%) | 113 (8.7) | 10 (12.5) | 103 (8.5) | 0.22 |
| Hypertension<sup>6</sup>, n (%) | 404 (31.3) | 33 (41.3) | 371 (30.6) | 0.05 |
| Asthma<sup>6</sup>, n (%) | 194 (15.0) | 13 (16.3) | 181 (14.9) | 0.75 |
| Cesarean delivery, n (%) | 400 (30.9) | 30 (37.5) | 370 (30.5) | 0.19 |
| Preterm delivery at gestational age <37 weeks, n (%) | 128 (9.9) | 11 (13.8) | 117 (9.7) | 0.23 |
| SARS-CoV-2 PCR tested during pregnancy<sup>7</sup>, n (%) | 1,109 (85.8) | 72 (90.0) | 1,037 (85.5) | 0.26 |
| Positive, n (% tested) | 64/1,109 (5.8) | 46/72 (63.9) | 18/1,037 (1.7) | <0.001 |
| Duration (in hours) between serology test and first positive PCR test<sup>8</sup>, median (IQR) | 0.3<br>(0, 345.3) | 75.4<br>(0, 659.3) | 0<br>(-0.1, 0.2) | 0.007 |
| Live-born infant, n (%) | 1,282 (99.2) | 79 (98.8) | 1,203 (99.2) | 0.51 |

**Footnotes:** <sup>1</sup>Seropositivity was based on either IgG or IgM level >0.48 arbitrary units.
<sup>2</sup>Difference in maternal age was tested using Mann-Whitney U test, differences in proportion for all other characteristics were tested using $\chi^2$ tests or Fisher's exact tests as appropriate. For race/ethnicity and pre-pregnancy BMI, difference was tested at each level of the characteristic (e.g. Black women compared to non-Black women). <sup>3</sup>Race/ethnicity was unknown for 2 seropositive and 26 seronegative women; race was abstracted from documentation at time of admission and in clinical practice is usually self-reported. <sup>4</sup>Health insurance was missing for 2
seropositive women; health insurance was abstracted from documentation in the medical record of the insurance used during admission for delivery; ‘Public payor’ includes Medicaid, Medicare and Veterans Affairs Community Care; ‘Uninsured’ includes self-pay and Charity Care. <sup>5</sup>Pre-pregnancy BMI was missing for 2 seropositive and 12 seronegative women; pre-pregnancy BMI was abstracted from documentation in the medical record or patient’s self-reported entry in birth registration. <sup>6</sup>Diagnoses were based on delivery admission International Classification of Diseases, 10<sup>th</sup> revision diagnosis codes for diabetes (O24, E08-E13, Z79.4), hypertension (O10, O11, O13-O16, I10-I13, I15) and asthma (J45). <sup>7</sup>Included tests done anytime during pregnancy up to discharge from delivery admission. <sup>8</sup>Difference calculated by subtracting the date and time sample for PCR was collected from the date and time sample for serology was collected. A negative number indicates PCR sample collected after serology sample. BMI, body mass index; IQR, interquartile range; PCR, polymerase chain reaction; SARS-CoV-2, severe acute respiratory syndrome coronavirus 2.

**Figure 1.**
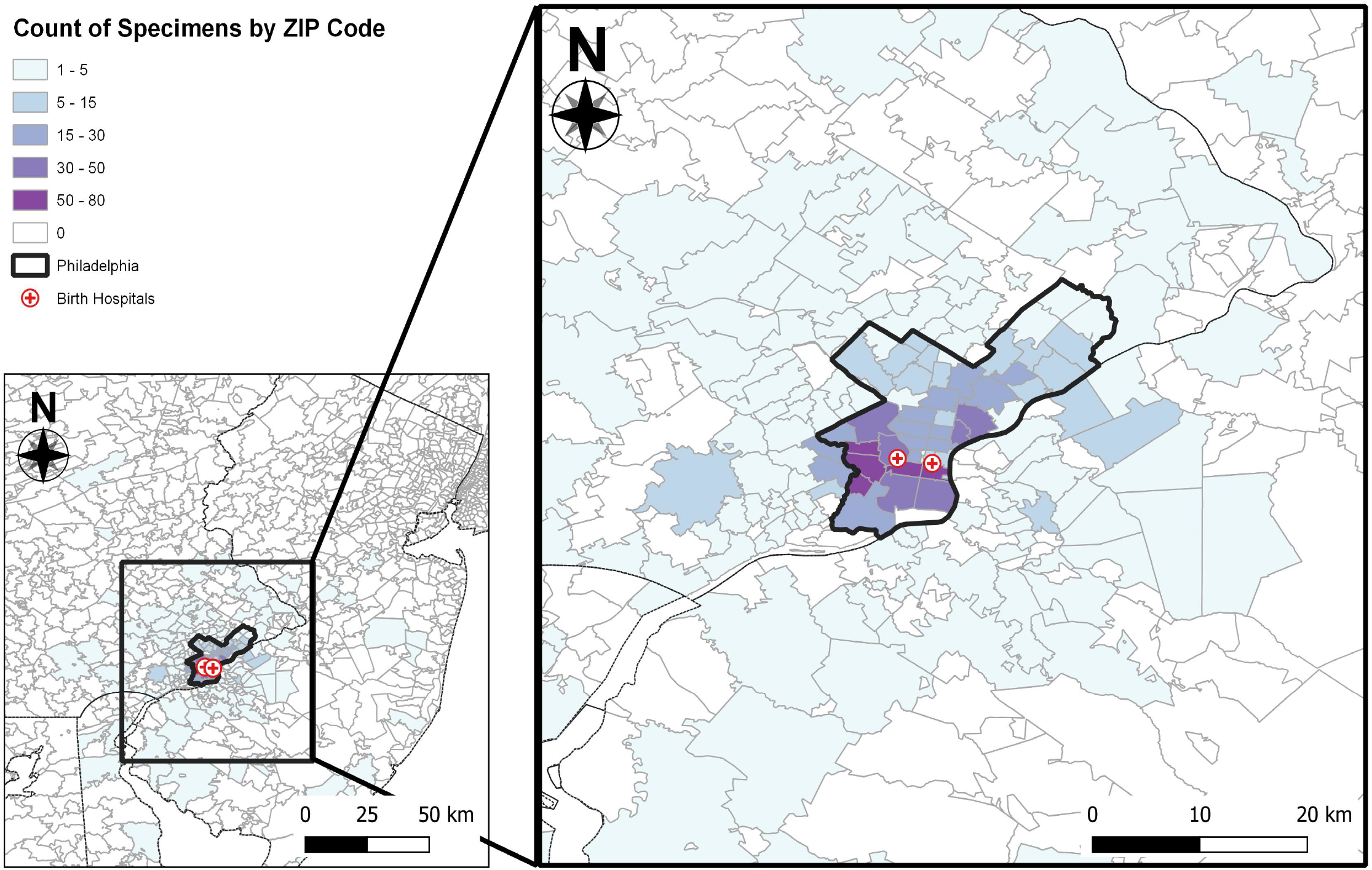
Geographical distribution of women tested for SARS-CoV-2 antibodies. Most serum specimens analyzed were from women living in areas within or immediately bordering the city of Philadelphia. Location of birth hospitals where serum samples were collected are shown as red crosses.

Our serological assay utilized a SARS-CoV-2 spike RBD antigen and modified ELISA protocol first described by Amanat *et al*. (6). We validated this serological assay by testing serum samples collected prior to the pandemic in 2019 from 834 individuals in the Penn Medicine Biobank and 15 individuals who recovered from confirmed coronavirus disease 19 (COVID-19) infections in 2020 (**Figure 2A-B**). All 15 serum samples from COVID-19 recovered donors contained high, but variable, levels of SARS-CoV-2 IgG (**Figure 2A**) and 10 of 15 samples contained detectable levels of SARS-CoV-2 IgM (**Figure 2B**). Conversely, only 5 of 834 samples collected before the pandemic contained SARS-CoV-2 IgG and only 4 of 834 samples contained SARS-CoV-2 IgM; none contained both IgG and IgM. Together, this indicates that there is an overall false positive rate ∼1% (9/834) in our serological assay. Consistent with our initial validation experiments, only 1 of 140 samples collected from pregnant women before the pandemic (from 2009-2012) possessed IgG or IgM SARS-CoV-2 antibodies (**Figure 2C-D**).

**Figure 2.**
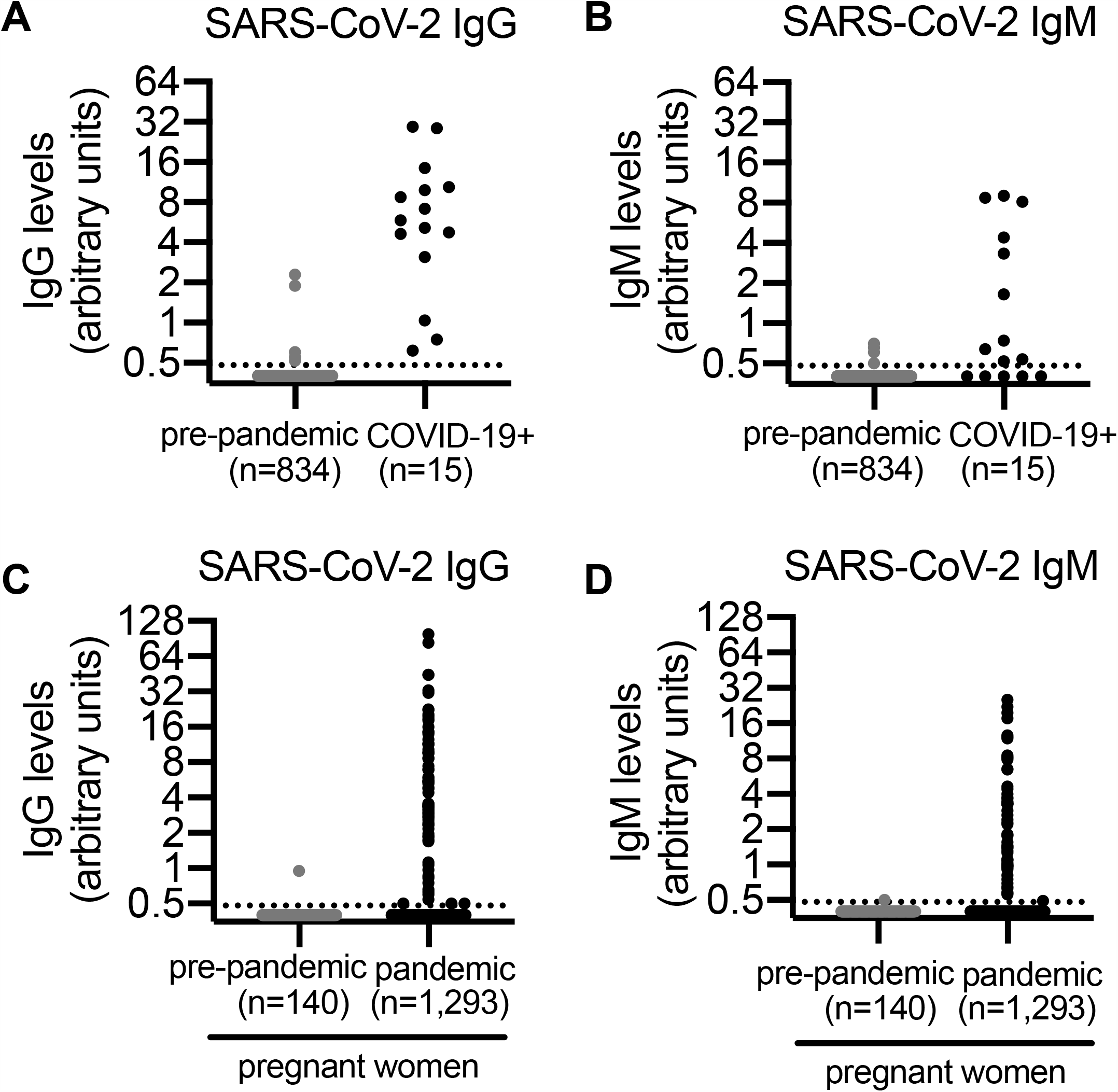
Serum SARS-CoV-2 antibody levels in COVID-19 pandemic and pre-pandemic individuals. (A-B) Relative levels of SARS-CoV-2 IgG (A) and IgM (A) in serum collected before the COVID-19 pandemic (n = 834) and serum collected from COVID-19 recovered donors (n = 15). (C-D) Relative levels of SARS-CoV-2 IgG (C) and IgM (D) in serum collected from pregnant women from 2009-2012 (n = 140) and from April 4-June 3, 2020 (n = 1,293). Dashed lines indicate 0.48 arbitrary units, which was used to distinguish positive versus negative samples (see Methods). Serum samples that were below the cutoff for seropositivity were assigned an antibody level of 0.40 arbitrary units.

We found that 80 of 1,293 (6.2%) pregnant women presenting for delivery from April 4 to June 3, 2020 possessed IgG or IgM SARS-CoV-2 antibodies (**Figure 2C-D**; p = 0.003 comparing samples from pre-pandemic and pandemic pregnant women). We identified 55 women with both SARS-CoV-2 IgG and IgM, 21 women with only SARS-CoV-2 IgG, and 4 women with only SARS-CoV-2 IgM (**Table 2**). SARS-CoV-2 antibody levels in samples from these women were variable (**Figure 2C-D)**, similar to what we found in samples from individuals recovering from confirmed SARS-CoV-2 infections (**Figure 2A-B**). The seroprevalence rate was not statistically different comparing women living within the city limits of Philadelphia (62/986, 6.3%) to those living in surrounding areas in Pennsylvania (12/191, 6.3%), or surrounding areas in New Jersey (5/107, 4.7%). In contrast, we observed significant race/ethnicity differences in seroprevalence rates with higher rates in Black/non-Hispanic (9.7%) and Hispanic/Latino (10.4%) women and lower rates in White/non-Hispanic (2.0%) and Asian (0.9%) women (**Table 1**).

**Table 2.** Relative levels of SARS-COV-2 IgG and IgM in serum collected from seropositive pregnant women (n = 80).

| **Patient Number** | **IgG levels (arbitrary units)** | **IgM levels (arbitrary units)** |
| --- | --- | --- |
| 1 | <0.20 | 0.57 |
| 2 | <0.20 | 0.99 |
| 3 | 0.29 | 0.75 |
| 4 | 0.31 | 1.09 |
| 5 | 0.50 | 0.21 |
| 6 | 0.50 | 1.04 |
| 7 | 0.50 | 2.28 |
| 8 | 0.54 | 0.35 |
| 9 | 0.60 | 0.65 |
| 10 | 0.63 | <0.20 |
| 11 | 0.66 | 1.01 |
| 12 | 0.75 | <0.20 |
| 13 | 0.83 | <0.20 |
| 14 | 0.86 | <0.20 |
| 15 | 0.97 | 0.43 |
| 16 | 1.07 | 0.49 |
| 17 | 1.12 | <0.20 |
| 18 | 1.70 | 0.56 |
| 19 | 1.75 | 0.69 |
| 20 | 1.85 | 0.33 |
| 21 | 1.87 | 0.29 |
| 22 | 1.97 | <0.20 |
| 23 | 2.16 | 0.30 |
| 24 | 2.34 | 0.36 |
| 25 | 2.35 | 1.78 |
| 26 | 2.45 | 1.44 |
| 27 | 2.58 | 0.98 |
| 28 | 2.73 | 0.30 |
| 29 | 2.75 | 2.22 |
| 30 | 2.85 | 0.91 |
| 31 | 2.94 | 0.39 |
| 32 | 2.99 | 2.90 |
| 33 | 3.12 | 1.10 |
| 34 | 3.20 | 0.28 |
| 35 | 3.32 | 0.21 |
| 36 | 3.34 | 1.11 |
| 37 | 3.38 | 1.40 |
| 38 | 3.42 | 0.58 |
| 39 | 3.46 | <0.20 |
| 40 | 3.54 | <0.20 |
| 41 | 3.55 | 1.40 |
| 42 | 4.40 | <0.20 |
| 43 | 4.75 | 0.96 |
| 44 | 4.88 | 0.94 |
| 45 | 5.25 | 3.37 |
| 46 | 5.43 | 2.41 |
| 47 | 5.46 | 1.77 |
| 48 | 5.50 | 12.25 |
| 49 | 5.59 | 3.43 |
| 50 | 5.87 | 6.52 |
| 51 | 5.97 | 3.28 |
| 52 | 6.91 | 6.44 |
| 53 | 6.94 | <0.20 |
| 54 | 7.40 | 3.27 |
| 55 | 8.60 | 3.71 |
| 56 | 8.67 | 0.98 |
| 57 | 9.56 | 12.60 |
| 58 | 9.59 | 1.26 |
| 59 | 9.80 | 1.32 |
| 60 | 10.23 | 4.40 |
| 61 | 11.28 | 25.33 |
| 62 | 11.47 | 0.81 |
| 63 | 11.52 | 7.90 |
| 64 | 12.45 | 4.63 |
| 65 | 13.97 | 1.45 |
| 66 | 14.44 | 1.56 |
| 67 | 14.46 | 8.29 |
| 68 | 15.99 | 2.64 |
| 69 | 18.10 | 0.63 |
| 70 | 18.76 | 3.99 |
| 71 | 19.56 | 1.82 |
| 72 | 20.47 | 19.70 |
| 73 | 22.32 | 3.32 |
| 74 | 22.52 | 8.49 |
| 75 | 31.26 | 3.67 |
| 76 | 32.85 | 22.02 |
| 77 | 44.26 | 17.59 |
| 78 | 44.46 | 3.27 |
| 79 | 83.77 | 8.07 |
| 80 | 99.10 | 11.87 |

NP swabs from 1,109 (85.8%) women were tested by SARS-CoV-2 PCR during the pregnancy or at the time of delivery. We found that 46 of 72 seropositive women who were NP tested had a SARS-CoV-2 positive PCR result, whereas only 18 of 1,037 seronegative women who were NP tested had a SARS-CoV-2 positive PCR result (**Table 1;** p <0.001). While all serum samples were collected during the delivery admission, NP samples were collected at variable times either during the delivery admission or earlier in the pregnancy, and therefore, further study will be required to evaluate the temporal relationship between SARS-CoV-2 seropositivity and PCR positivity in pregnant women.

Large-scale serology testing is critical for estimating how many individuals have been infected during the COVID-19 pandemic. Due to widely-imposed social distancing requirements, and to decreases in on-site, discretionary medical care, it is currently difficult to collect serum for population-wide serological testing. The vast majority of pregnant women, however, continue to have multiple interactions with the medical system for prenatal care and for delivery during this pandemic, and therefore represent a unique population to assess SARS-CoV-2 immunity within a community. Our data suggest that ∼5.2% (6.2% minus ∼1% false positive rate) of parturient women in Philadelphia from April 4 to June 3, 2020 were previously exposed to SARS-CoV-2. As of June 3, 2020, there were 23,160 confirmed cases of COVID-19 in the city of Philadelphia (7), which has a population size of nearly 1.6 million people, suggesting an infection rate of approximately 1.4%. Serologic studies may provide a more accurate means of assessing population exposure to SARS-CoV-2 by identifying asymptomatic or minimally symptomatic as well as symptomatic infections. Further studies are needed to determine how the immune status of pregnant women compares to that of the general population. For example, parturient women may not represent individuals of different ages within the general population and women and men might mount different antibody responses upon infection with SARS-CoV-2 (8).

Pregnant women of all demographics continue to seek medical care during the COVID-19 pandemic. Therefore, parturient women represent a unique population to assess differences in SARS-CoV-2 exposures in diverse populations. Our finding that Black/non-Hispanic and Hispanic/Latino women have higher SARS-CoV-2 seroprevalence rates relative to women of other races suggest that there are race/ethnicity differences in SARS-CoV-2 exposures in Philadelphia and surrounding areas. Identification of factors that contribute to such differences in exposure to SARS-CoV-2 may inform public health measures aimed at preventing further infections.

Prior perinatal COVID-19 studies have primarily focused on virus detection (i.e. nucleic acid testing) in pregnant women and have not evaluated immunity (9–17). Two published studies to date have assessed SARS-COV-2 serology in pregnant women with active disease. A study of 6 parturient women in Wuhan, China with confirmed COVID-19 found all 6 women had elevated levels of SARS-CoV-2 IgG and IgM (18). A case report from Peru detailed a symptomatic pregnant woman with positive PCR testing and negative serology at presentation, who developed severe respiratory failure necessitating delivery; her IgM and IgG turned positive 4 days after delivery (9 days after symptom onset) (19). Beyond describing individual response to infection, SARS-CoV-2 serological testing among pregnant women will be increasingly important for perinatal disease risk management, as well as for optimizing vaccine strategies when vaccines become available. Additional studies will be needed to address the impact of maternal infection on neonatal immunity, and to determine those factors that may contribute to observed disparities in exposure to SARS-CoV-2.

## Data Availability

All data are included in the manuscript.

## Acknowledgments

We thank all members of the Wherry Lab and the Penn COVID-19 Sample Processing Unit for sample procurement, processing and logistics. We thank the staff of the Penn Medicine Biobank. We thank Florian Krammer (Mt. Sinai) for sending us the SARS-CoV-2 spike RBD expression plasmids. We thank Dr. Steven Melly (Drexel University) for his assistance in geographic analyses.

## Funding

This work was supported by institutional funds from the University of Pennsylvania and an NIH grant AI082630 (to E.J.W.). We thank Jeffrey Lurie and we thank Joel Embiid, Josh Harris, David Blitzer for philanthropic support. E.J.W. is supported by the Parker Institute for Cancer Immunotherapy which supports the cancer immunology program at UPenn.

## Author contributions

DDF conceptualized and designed the study, collected data, drafted the initial manuscript, and revised the manuscript. SG led the serological experiments, collected data, and revised the manuscript. MBD designed the data collection instruments, collected data, carried out the analyses, and revised the manuscript. SM conceptualized and designed the study, designed the data collection instruments, carried out the analyses, and revised the manuscript. MRP collected data and revised the manuscript. ECW collected data and revised the manuscript. JSG conceptualized and designed the study, and revised the manuscript. CPA completed serological assays, analyzed data, and revised the manuscript. MJB completed serological assays, analyzed data, and revised the manuscript. MW completed serological assays, analyzed data, and revised the manuscript. ECG completed serological assays, analyzed data, and revised the manuscript. EMA completed serological assays, analyzed data, and revised the manuscript. ARG obtained and proceeded samples from recovered donors. JK obtained and proceeded samples from recovered donors. NH obtained and proceeded samples from recovered donors. AP obtained and proceeded samples from recovered donors. JD obtained and proceeded samples from recovered donors. OK designed and established recovered donor cohort. DM processed and characterized samples from recovered donors. AB oversaw acquisition, processing, and characterization of samples from recovered donors. LAV designed and established recovered donor cohort. JW supervised recruitment of participants in PMBB and identification of samples for serology testing. AV analyzed demographic data of PMBB participants. RL provided samples for the pre pandemic pregnant controls. JSM provided statistical advice, performed statistical analyses, and revised the paper. DJR provided input on the use of PMBB controls and revised the manuscript. MAE provided input and samples for the pre-pandemic pregnant controls and revised the manuscript. EJW designed, established, and oversaw healthy donor cohort studies and made revisions to the manuscript. KMP conceptualized and designed the study, coordinated and supervised data collection, and revised the manuscript. SEH conceptualized and designed the study, coordinated and supervised serological studies, and revised the manuscript.

## Competing interests

SEH has received consultancy fee from Sanofi Pasteur, Lumen, Novavax, and Merck for work unrelated to this report. EJW is a member of the Parker Institute for Cancer Immunotherapy. EJW has consulting agreements with and/or is on the scientific advisory board for Merck, Roche, Pieris, Elstar, and Surface Oncology. EJW is a founder of Surface Oncology and Arsenal Biosciences. EJW has a patent licensing agreement on the PD-1 pathway with Roche/Genentech. All other authors declare no competing interests related to this work.

## Data and materials availability

All data are included in the manuscript.

## Supplementary Materials

Materials and Methods

## Notes

### Clinical Trial

residual samples used, no trial ID

### Author Declarations

The Institutional Review Board at the University of Pennsylvania approved this study with waiver of consent.

